# Changes in disability status and oral healthcare affordability among working-age Australians

**DOI:** 10.64898/2026.07.21.26358621

**Authors:** Upul Cooray, Gagandeep Kaur, Saman Khalatbari-Soltani, Barbara Janssens, George Disney, Ruby Cole, Ankur Singh

## Abstract

**Importance:** Oral healthcare is often financed outside universal medical coverage, leaving working-age adults exposed to out-of-pocket costs. People with long-term disability may face added financial, physical, and service barriers to care, but longitudinal evidence on disability and oral healthcare unaffordability is limited.

**Objective:** To estimate the effect of time-varying long-term disability on oral healthcare unaffordability among working-age adults in Australia.

**Design:** Longitudinal cohort study using Household, Income and Labour Dynamics in Australia survey data from waves 18 to 22 (2018-2022), analysed with targeted maximum likelihood estimation for longitudinal modified treatment policies.

**Setting:** Nationally representative household panel survey in Australia.

**Participants:** Adults aged 25 to 65 years at wave 18 who could validly contribute to the longitudinal analysis (identified using HILDA longitudinal weights) and had complete baseline covariate data.

**Exposure:** Time-varying self-reported disability at waves 18 to 21 (2018-2022), defined as any long-term health condition, impairment, or disability restricting everyday activities and lasting, or likely to last, for at least 6 months. Hypothetical interventions comprised 50% and 25% reductions in the odds of disability at each wave, and deterministic sustained disability and no disability regimes.

**Main Outcome and Measure:** Self-reported avoidance of dental treatment because of cost at wave 22 (2022).

**Results:** The analytic sample included 9635 adults; 4901 (50.9%) were female, mean age was 44 (SD=12) years, and 2419 (25.1%) reported disability at baseline. A total of 399 participants (4.1%) reported oral healthcare unaffordable at wave 22 follow-up. Compared with the natural course, sustained disability increased the risk of unaffordability (risk ratio [RR], 1.60; 95% CI, 1.15-2.22). No disability at any time point reduced the risk (RR, 0.59; 95% CI, 0.45-0.77). Reducing the odds of disability by 50% and 25% also reduced the risk of oral healthcare unaffordability by 28% (RR, 0.72; 95% CI, 0.65-0.81) and 17% (RR, 0.83; 95% CI, 0.78-0.89), respectively.

**Conclusions and Relevance:** Under the study assumptions, long-term disability was estimated to increase experienced unaffordability of oral healthcare among working-age Australians. Population level policy responses should address both the upstream conditions that shape disability trajectories and the downstream exclusion of adult dental care from routine financial protection.

**Key Points:** *Question:* What is the estimated effect of time-varying disability status on oral healthcare affordability among working-age adults in Australia?

*Findings:* In this longitudinal cohort analysis of 9635 working-age adults, reducing the odds of disability by 50% or 25% was associated with lower risk of finding oral healthcare unaffordable at follow-up, while sustained disability was associated with higher risk.

*Meaning:* The findings support paired policy attention to disability prevention and oral healthcare financing protections for working-age adults with disability.

## Introduction

Service coverage and financial protection are central to universal health coverage, as access to necessary healthcare should not depend on an individual’s capacity to pay.^1^ Oral healthcare remains a common exception to this principle. Across health systems, oral healthcare services are often financed through private payment or partial insurance rather than comprehensive public coverage, and cost is a frequent reason for delayed or forgone oral healthcare.^1–3^ In Australia, most oral health services are delivered in private settings, Medicare does not cover adult oral healthcare, and public oral health services for adults are eligibility restricted.^4,5^ This system makes oral healthcare affordability a persistent policy problem, particularly for working-age adults, who fall outside age-based concessions available to younger and some older Australians.

Long-term disability can increase the need for oral health and other healthcare services while reducing income, employment security, transport options, and the ability to locate services that can accommodate physical, cognitive, or psychosocial needs.^6–9^ For working age adults, these barriers can arise before eligibility for age-based services subsidies and retirement income support, leaving this group disproportionately affected by the cost of services. Cost is therefore an important part of the pathway from disability to unmet oral healthcare need and poor oral health status. Moreover, cost is the main pathway measured directly in many surveys related to healthcare accessibility, including the outcome used in this study. Prior studies show that adults with disability are more likely than adults without disability to delay or forgo healthcare, including oral healthcare, because of cost.^6,8,9^

However, much of the evidence is cross-sectional or treats disability as a fixed baseline attribute. Those designs cannot capture how disability and associated factors such as employment and income change together over time. They also cannot separate the effect of disability on affordability from the reverse pathway, in which disability changes employment or income, which then affects whether disability is recorded again at a later wave. When a covariate measured after baseline predicts subsequent exposure and independently predicts the outcome, while also potentially being affected by prior exposure, it should be treated as time-varying confounding.^10^ Standard regression adjustment cannot handle this time-varying confounding structure correctly.^11^ Furthermore, treating disability as fixed baseline status compounds this issue. For example, if disability is present at baseline but resolves partway through follow-up and is coded as ‘disability’ for the whole study period, the time after it resolves is misclassified as exposed time. Outcomes from that unexposed (‘no disability’) period are pooled into the disability exposed group biasing the estimated effect of disability on affordability.^12,13^

This study estimated the effect of changes in disability status on experienced unaffordable oral healthcare among working-age Australian adults. We used longitudinal modified treatment policies to compare the observed natural course with hypothetical interventions that reduced the odds of experienced disability at each wave, and with counterfactual sustained disability and no disability regimes.

## Methods

### Study design and participants

We conducted a longitudinal cohort analysis using the Household, Income and Labour Dynamics in Australia (HILDA) survey, a nationally representative household panel study.^14^ The primary analysis used waves 18 to 22 (2018-2022). Participants were eligible if they were aged 25 to 65 years at wave 18, had non-zero longitudinal survey weights to ensure that each participant could validly contribute to the longitudinal analysis, and had complete data on baseline covariates. The participant selection flowchart is shown in Figure 1.

**Figure 1.**
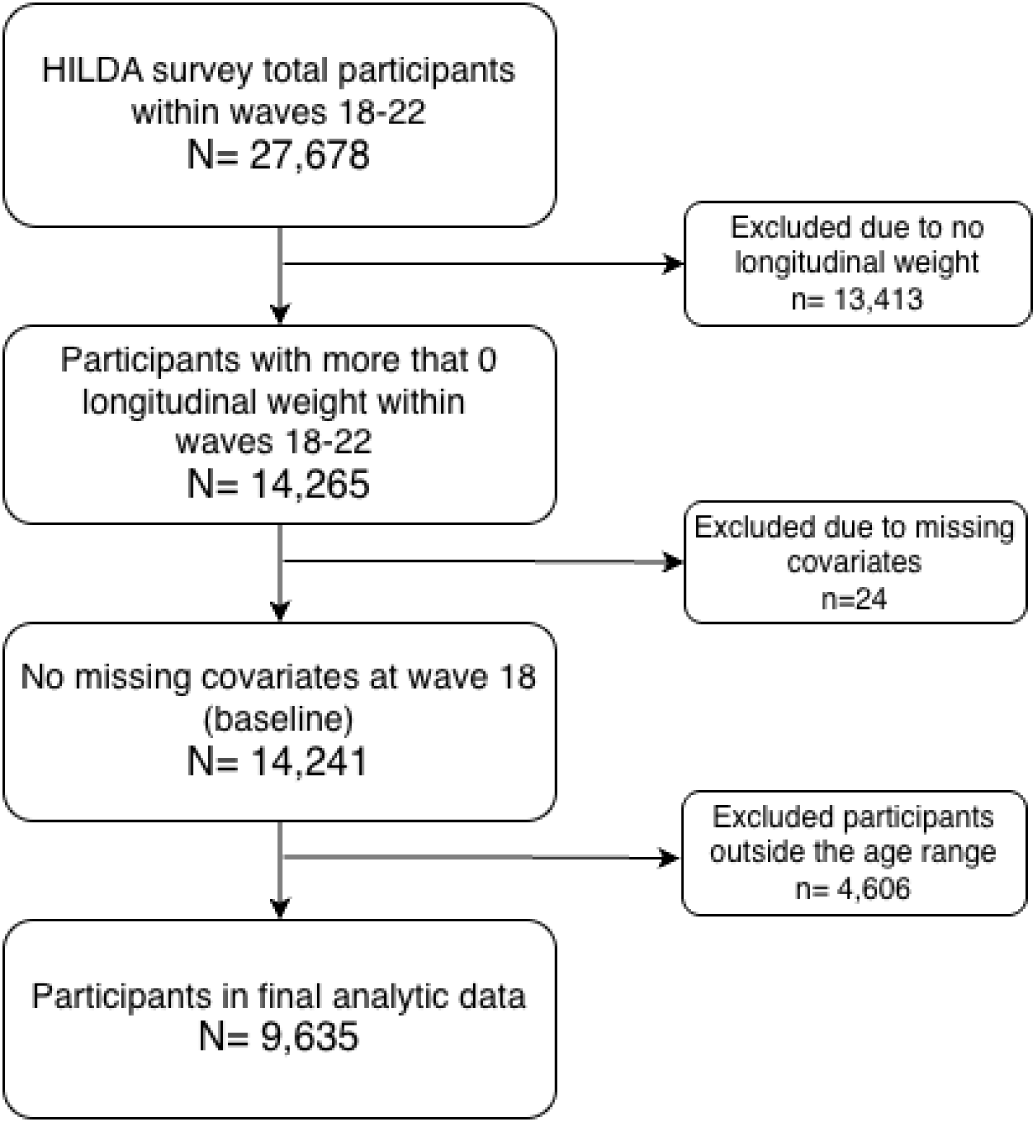
**Participant selection flowchart**

### Exposure and hypothetical interventions

The exposure was self-reported disability at each wave from waves 18 to 21. HILDA asks participants, “do you have any long-term health condition, impairment or disability that restricts you in your everyday activities, and has lasted or is likely to last, for six months or more?”.

People who answered “Yes” were classified as having disability at that wave. The wording combines current restriction with expected duration; therefore, each wave was treated as a time-specific report of long-term activity restriction, not as a direct measure of exactly 12 months of disability.

We evaluated four hypothetical intervention regimes. Two were modified treatment policies that randomly reduced individuals’ odds of experienced disability at each wave by 50% and 25%, implemented as Incremental Propensity Score Interventions (IPSI).^15^ For example, under the 50% reduction policy, a participant’s probability of reporting disability at a given wave was shifted downward conditional on their measured history, rather than being set to zero for all participants. We also implemented two deterministic contrasts: sustained disability experience at every exposure wave and no disability at any exposure wave. The IPSI scenarios were the primary policy relevant contrasts because they preserve the observed exposure and covariate history at each time point, which keeps the shifted exposure distribution close to the empirical support of the data. This makes IPSI scenarios less prone to practical positivity violations than the deterministic extremes, which can require support for combinations of exposure and covariate history that are rare or absent in the observed data.^12,16^ Supplementary Figure S1 illustrates observed and shifted disability trajectories in a random sample of participants.

### Outcome

Experienced unaffordable oral healthcare was the outcome. In the HILDA survey unaffordable oral healthcare is measured every 5 years. The main analysis used the latest outcome measured at wave 22. People who found oral healthcare unaffordable were identified if they reported avoiding needed dental treatment and reported cost as the reason. This outcome captures experienced unaffordable oral healthcare among people who perceived a need using HILDA questions “Do you (and your family) have dental treatment when needed?”. And, for those who responded “No”, “Is that because you cannot afford it?”. It does not capture unmet normative dental need among people who did not perceive a need for treatment.

### Covariates and censoring

The adjustment set was informed by the directed acyclic graph (DAG; Figure 2). Baseline covariates were age (continuous - in years), sex (female/ male), highest educational attainment (bachelor or above/ high school or diploma/ less than high school), and country of birth (Australia/ New Zealand/ United Kingdom/ Other). Time-varying covariates were equivalised annual household income (continuous - in Australian dollars adjusted for household size using modified OECD equivalence scale)^17^, employment status (employed/ unemployed/ not in the labour force), remoteness area (major city/ inner regional/ outer regional/ remote), and marital status (married/ living with partner/ separated or widowed/ never married). These covariates were treated as time-varying confounders because they may affect subsequent disability and oral healthcare affordability, while also being affected by earlier disability.^18^ Right censoring due to dropout was handled within the longitudinal targeted maximum likelihood estimation framework.^19^ The flow of participant numbers after each follow-up wave is shown in Supplementary Table S1.

**Figure 2.**
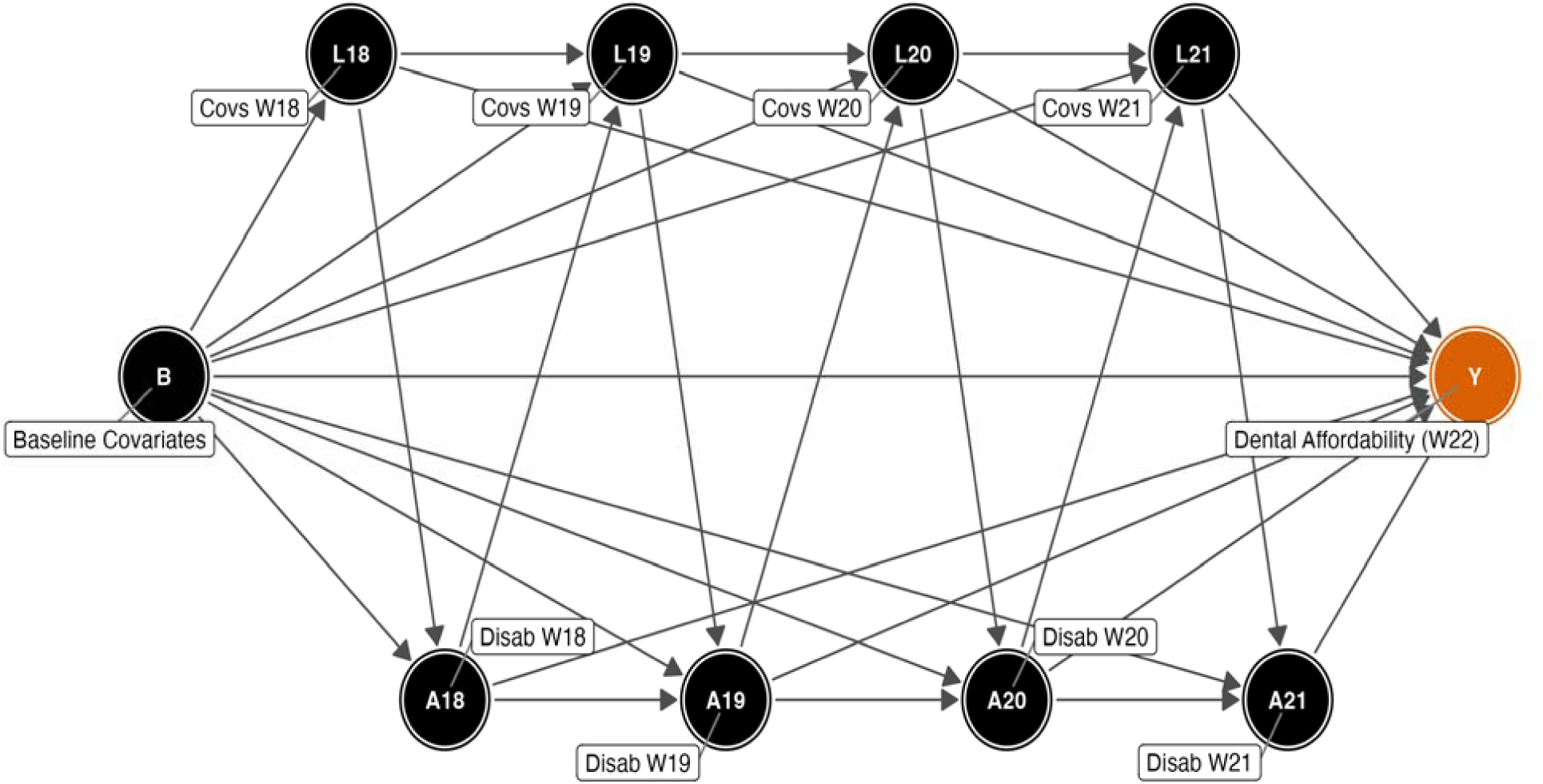
Simplified directed acyclic graph (DAG) Note: The DAG shows the assumed causal structure across waves 18 to 22. B denotes baseline covariates (age, sex, education, and country of birth). L18-L21 denote time-varying confounders measured at each exposure wave (equivalised household income, employment status, remoteness, and marital status). A18-A21 denote disability status (the exposure) at waves 18 to 21. Y denotes the outcome, oral healthcare unaffordability, measured at wave 22. Arrows represent assumed direct effects: each variable may affect later confounders, later disability, and the outcome, and earlier disability may affect later confounders (time-varying confounding affected by prior exposure).

### Statistical analysis

For each hypothetical intervention regime, we targeted the population mean counterfactual outcome and estimated risk ratios (RR) and risk differences (RD) relative to the observed natural course. Estimation used targeted maximum likelihood estimation for longitudinal modified treatment policies using a Super Learner ensemble that included generalised linear models, generalised additive models, extreme gradient boosting, and neural networks.^12,20,21^ This approach allows flexible nuisance parameter estimation (i.e. underlying prediction models for the outcome, the exposure, and censoring which are needed for estimation but are not the final effect of interest) while accounting for time-varying confounding and censoring. To guard against overfitting and support valid confidence intervals, all models were cross-validated using fivefold cross-fitting at the targeted maximum likelihood estimation level and twofold inner cross validation for Super Learner training.^22^ Causal identification depends on consistency, positivity, and conditional exchangeability, meaning no unmeasured common causes of disability and oral healthcare affordability conditional on the measured covariate history.^23^

Missingness values were handled using multivariate imputation by chained equations with 10 imputed data sets and the estimates were pooled using Rubin rules.^24,25^ HILDA survey weights were normalised and used in estimation. The distribution of missingness across study variables is shown in Supplementary Figure S2.

We calculated E-values for point estimates and the confidence interval limit closest to the null. The E-value indicates the minimum strength of effect, on the risk ratio scale, that an unmeasured confounder would need to have with both disability and oral healthcare affordability, conditional on measured covariates, to explain away the observed estimate.^26,27^ Furthermore, two sensitivity analyses were conducted: an incidence cohort analysis restricted to participants who did not report oral healthcare as unaffordable at baseline, and a pre-COVID-19 pandemic analysis using waves 14 to 18. The incidence cohort analysis estimates reflect new rather than pre-existing affordability; the pre-pandemic analysis tested whether the findings were robust to the service disruptions and economic shocks of the COVID-19 period.

All the analyses were conducted using R version *4.5.2* in Positron version *2026.06.0* for *aarch64-apple-darwin20*. Targeted maximum likelihood estimation for longitudinal modified treatment policies was conducted using *lmtp and SuperLearner* R packages.^16^ All R code used to generate the main results is publicly available at https://github.com/upulcooray/HILDA_Disability. Reporting of this study conforms to the STROBE guidelines.

## Results

### Participant characteristics

The analytic sample included 9635 working-age adults (Figure 1). A total of 399 participants (4.1%) reported oral healthcare unaffordable at wave 22. At baseline, 2419 participants (25.1%) had disability; this increased to 2632 (28.2%) at wave 21. Across all waves (18 to 21), 5598 participants (58.1%) never had disability and 1271 (13.2%) had disability at every exposure wave.

Across waves, people with disability differed markedly from those who did not (Table 1). At wave 18, adults with disability had lower equivalised household income than those without disability (mean, $48,199 vs $63,976). They were less likely to have a bachelor’s degree or higher qualification (23% vs 41%) and more likely to have less than high school education (26% vs 11%). People with disability were also more likely to be not in the labour force (44% vs 13%). These differences were consistent across waves 18 to 21.

**Table 1.** Participant characteristics by disability status at each wave, waves 18-21.

| Variable | Wave 18<br>Disability |  | Wave 19<br>Disability |  | Wave 20<br>Disability |  | Wave 21<br>Disability |  |
| --- | --- | --- | --- | --- | --- | --- | --- | --- |
|  | No | Yes | No | Yes | No | Yes | No | Yes |
| N | 7,214 | 2,419 | 7,053 | 2,427 | 6,893 | 2,490 | 6,687 | 2,632 |
| Age, mean (SD) | 42 (11) | 49 (12) | 43 (11) | 49 (12) | 42 (11) | 49 (12) | 43 (11) | 48 (12) |
| Sex- female | 50.4% | 52.4% | 50.8% | 52.1% | 50.8% | 53.2% | 50.1% | 55.3% |
| Highest education |  |  |  |  |  |  |  |  |
| Bachelor or above | 41.1% | 23.4% | 40.9% | 23.4% | 40.9% | 23.8% | 40.8% | 25.2% |
| High school (HS) | 47.6% | 50.8% | 47.9% | 50.9% | 48.5% | 49.6% | 47.9% | 50.7% |
| Less than HS | 11.3% | 25.8% | 11.2% | 25.7% | 10.6% | 26.7% | 11.2% | 24.1% |
| Equivalised household income*, mean (SD) | 63,976 (35,499) | 48,199 (30,126) | 66,553 (40,381) | 52,736 (35,200) | 68,712 (40,636) | 52,914 (31,002) | 72,134 (44,883) | 55,118 (33,545) |
| Employment Status |  |  |  |  |  |  |  |  |
| Employed | 84.9% | 51.7% | 84.4% | 51.5% | 83.6% | 49.1% | 83.5% | 50.9% |
| Not in labour force | 12.9% | 43.8% | 13.8% | 44.0% | 13.7% | 46.7% | 14.7% | 44.1% |
| Unemployed | 2.2% | 4.5% | 1.8% | 4.5% | 2.7% | 4.3% | 1.8% | 5.0% |
| Remoteness area |  |  |  |  |  |  |  |  |
| Major City | 71.6% | 63.0% | 70.3% | 63.6% | 70.5% | 62.2% | 70.5% | 60.7% |
| Inner Regional Australia | 19.5% | 26.0% | 20.2% | 26.5% | 21.0% | 27.3% | 21.2% | 27.6% |
| Outer Regional Australia | 7.6% | 9.6% | 8.2% | 8.5% | 7.4% | 9.1% | 7.1% | 10.5% |
| Remote Australia | 1.3% | 1.4% | 1.3% | 1.3% | 1.1% | 1.4% | 1.2% | 1.3% |
| Marital Status |  |  |  |  |  |  |  |  |
| Married | 58.8% | 47.6% | 60.1% | 49.0% | 60.3% | 48.9% | 61.6% | 48.4% |
| Living with partner | 15.6% | 12.3% | 15.6% | 12.4% | 14.8% | 12.5% | 14.5% | 11.4% |
| Never married | 16.9% | 21.8% | 15.3% | 20.1% | 15.2% | 19.6% | 13.8% | 20.7% |
| Separated or widowed | 8.7% | 18.3% | 9.0% | 18.5% | 9.7% | 19.0% | 10.1% | 19.5% |
| Country of birth |  |  |  |  |  |  |  |  |
| Australia | 65.1% | 75.0% | 66.1% | 74.7% | 65.7% | 76.2% | 67.0% | 72.9% |
| New Zealand | 4.1% | 1.7% | 3.7% | 2.0% | 3.8% | 1.6% | 2.9% | 4.0% |
| UK | 4.7% | 4.8% | 4.4% | 5.0% | 4.3% | 5.2% | 4.3% | 5.1% |
| Other | 26.1% | 18.6% | 25.8% | 18.3% | 26.2% | 17.0% | 25.7% | 18.0% |

Note: Survey weighted percentages and means (with standard deviations, SD) are shown. Disability is self-reported, defined as any long-term health condition, impairment, or disability that restricts everyday activities and has lasted, or is likely to last, for at least 6 months.
\*Equivalised household income is annual household disposable income adjusted for household size and composition using the modified OECD equivalence scale<sup>17</sup>, expressed in Australian dollars.

### Estimated effects of disability interventions

Under the observed natural course of disability among participants, the estimated risk of experienced unaffordable oral healthcare was 0.0489 (95% CI, 0.0431 to 0.0548), i.e. 4.9%. Compared with the natural course, modified treatment policy scenarios produced graded improvements in affordability. A 50% reduction in the odds of disability reduced experienced unaffordable oral healthcare by 28% (RR= 0.72 [95% CI, 0.65-0.81]; RD= −0.013 [95% CI, - 0.018 to −0.009]). A 25% reduction also reduced risk, with an effect size of 17% (RR= 0.83 [95% CI, 0.78-0.89]; RD= −0.008 [95% CI, −0.011 to −0.005]). Deterministic interventions of sustained disability at every exposure wave increased experienced unaffordable oral healthcare by 60% (RR= 1.60 [95% CI, 1.15-2.22]; RD= 0.029 [95% CI, 0.003-0.056]). No disability at any exposure wave reduced experienced unaffordable oral healthcare risk by 41% (RR= 0.59 [95% CI, 0.45-0.77]; RD= −0.020 [95% CI, −0.028 to −0.012]). Calculated E-values suggested that moderately strong unmeasured confounding would be required to explain away the observed effect estimates. Table 2 summarises the results of the main analysis with their associated E-values.

**Table 2.** Estimated effects of disability interventions on oral healthcare unaffordability.

| **Intervention scenario** | **Risk ratio (95% CI)** | **Risk difference (95% CI)** | **E-value (CI limit)** |
| --- | --- | --- | --- |
| 50% reduction in odds of disability | 0.72 (0.65, 0.81) | -0.013 (-0.018, -0.009) | 2.10 (1.77) |
| 25% reduction in odds of disability | 0.83 (0.78, 0.89) | -0.008 (-0.011, -0.005) | 1.69 (1.51) |
| Sustained disability at each wave | 1.60 (1.15, 2.22) | 0.029 (0.003, 0.056) | 2.58 (1.57) |
| No disability at any wave | 0.59 (0.45, 0.77) | -0.020 (-0.028, -0.012) | 2.79 (1.93) |

Note: Risk ratios and risk differences compare each intervention scenario with the observed natural course. Negative risk differences indicate lower absolute risk of oral healthcare unaffordability. CI indicates confidence interval. The E-value indicates the minimum strength of association, on the risk ratio scale, that an unmeasured confounder would need with both disability and oral healthcare unaffordability, conditional on measured covariates, to explain away the observed estimate; the value in parentheses is the corresponding E-value for the confidence interval limit closest to the null.

### Sensitivity analyses results

In the incidence cohort analysis, the deterministic sustained disability estimate was elevated but imprecise (RR, 1.43; 95% CI, 0.89-2.30). The no disability regime and odds reduction policies remained consistent with lower risk (no disability: RR= 0.52 [95% CI, 0.40-0.67]; 50% reduction: RR= 0.71 [95% CI, 0.63-0.81]; 25% reduction: RR= 0.85 [95% CI, 0.79-0.91]). The pre-pandemic waves 14 to 18 analysis produced estimates in the same direction as the primary analysis. All the results from sensitivity analyses are reported in Supplementary Tables S2 and S3.

## Discussion

In this nationally representative longitudinal cohort of working-age Australians, hypothetical intervention policies showed a graded pattern whereby a 50% reduction in the odds of disability produced a larger effect than a 25% reduction. Sustained long-term disability was estimated to increase the risk of unaffordable oral healthcare, while no disability was estimated to reduce the risk. The results extend prior evidence that adults with disability are more likely to delay or forgo care because of cost.^6,8,9^ Earlier studies have mainly described differences at a single time point or by baseline disability status. This study adds a longitudinal estimand that quantifies how oral healthcare affordability would change if the distribution of disability impact over time shifted. This is an important consideration, as disability can affect income, employment, household circumstances, and place of residence, which can in turn affect future disability and the ability to pay for dental care.^6,28^

Several mechanisms may explain the estimated effect. Disability may reduce earnings or labour-force participation, increasing sensitivity to out-of-pocket dental costs. It may also increase the cost due to care complexity, transport needs, reliance on carers, or the difficulty of finding services able to accommodate physical, cognitive, or psychosocial needs.^6–9^ Our modified treatment policies were designed to estimate how oral healthcare affordability might change under population level shifts in the probability of reporting activity limiting long-term conditions. Such shifts could arise from chronic disease prevention, rehabilitation, workplace accommodation, social protection, accessible environments, and timely support services.^29,30^ These upstream strategies are complementary to, not substitutes for, oral healthcare financing reforms. This distinction is important in the Australian policy setting. Most adult oral healthcare is outside routine Medicare coverage, and access to public oral healthcare varies by jurisdiction and eligibility.^4,5^ Some people with disability may qualify for concession linked public oral health services, but eligibility does not remove barriers related to disability and other indirect costs associated with access to services.^31^ Furthermore, there could be a crowding-out effect on oral health when limited household resources are absorbed by the direct and indirect costs of managing a function limiting condition, more deferrable out-of-pocket spending such as oral healthcare is deprioritised against more pressing competing demands.^32^ The findings therefore support two linked policy responses: reducing preventable activity limitations where possible, alongside improving support for people with activity limitations and improving financial protection for oral healthcare among adults already living with disability.

These implications must be interpreted considering several study limitations, which we attempted to minimise through robust study design and analytical choices. First, the outcome captures avoidance of oral healthcare that people perceived they needed but avoided because of cost, hence, the estimates should be interpreted as effects on experienced affordability issue rather than on all forms of unmet oral health need. People who did not perceive or report a dental need could still have unmet clinical need, which would tend to underestimate the true burden.

Because both the disability exposure and the affordability outcome were self-reported, dependent measurement error is also possible and could bias estimates in either direction. Second, conditional exchangeability cannot be verified due to unmeasured confounders; however, we adjusted for a comprehensive set of time-varying socioeconomic factors and calculated E-values to quantify the strength of unmeasured confounding required to explain the observed effect estimates.^26^ The resulting E-values indicate that moderately strong unmeasured confounding would be required to explain away our estimates, suggesting that the observed associations are relatively robust. Third, the broad HILDA disability item combines current restriction with expected duration and does not distinguish impairment type or severity. Our modified treatment policies shift the population distribution of this composite item rather than resolve its heterogeneity, so the estimates should be read as responses to a population level shift in the odds of reporting an activity limiting condition rather than to changes in any specific type, severity, or duration of impairment.^15^ Finally, deterministic counterfactual extremes such as sustained disability may be unrealistic for progressive conditions, which is why we focused on exposure shifts that respect individual covariate histories.

Despite these limitations, the study has several methodological and structural strengths. The primary strength is our longitudinal design using five waves of a nationally representative cohort, which allowed disability and socioeconomic circumstances to vary dynamically over time. We addressed the resulting time-varying confounding and right censoring using targeted maximum likelihood estimation for modified treatment policies; standard regression models fail to account for these feedback loops without introducing overadjustment or collider bias.^33^ Furthermore, the use of modified treatment policies provided practically achievable, policy relevant contrasts that preserve the observed data distribution. Because the affordability measure asked specifically whether unmet need was due to cost rather than to other reasons, the outcome isolates the economic barrier from non-financial factors such as dental anxiety, yielding a clear estimand of affordability. The consistency of these estimates across our pre-pandemic sensitivity analysis indicates that the primary findings were not driven by COVID-19 period disruptions.

In conclusion, disability was estimated to reduce affordability of oral healthcare among working-age Australians. Partial reductions in the odds of disability were associated with lower risk of experienced unaffordable oral healthcare in a dose responsive manner. Policies improving disability prevention, management, accommodation, and social support should be considered alongside reforms that reduce out-of-pocket dental costs for adults with disability.

## Declarations

### Ethics Approval and Consent to Participate

The HILDA Survey is approved by the Human Research Ethics Committee of The University of Melbourne. This study used deidentified secondary data and did not require additional ethical approval.

## Data Availability Statement

The HILDA survey data used in this study are available to approved researchers through a restricted access data licence from the Melbourne Institute, University of Melbourne. Analytical code used in this study is available at https://github.com/upulcooray/HILDA_Disability.

## Author Contributions

Conceptualization: UC, AS, GD, GK, BJ, SKS. Methodology: UC, AS. Formal analysis: UC, GK. Writing - original draft: UC, AS, GK, RC. Writing - review and editing: BJ, SKS, GD, RC, AS.

## Data Availability

https://melbourneinstitute.unimelb.edu.au/hilda/for-data-users

https://github.com/upulcooray/HILDA_Disability

## Acknowledgement

This paper uses unit record data from the HILDA Survey. The HILDA Survey was initiated and is funded by the Australian Government Department of Social Services and is managed by the Melbourne Institute of Applied Economic and Social Research.

## Use of Generative AI

Microsoft Copilot was used to assist with manuscript editing and English grammar. The authors reviewed and revised the final manuscript content.

