## Supplementary material for "Changes in disability status and oral healthcare affordability among working-age Australians"

**eFigure S1. Observed and shifted disability trajectories in a random sample of participants, under the natural course and the 50% and 25% odds-reduction (IPSI) policies.**

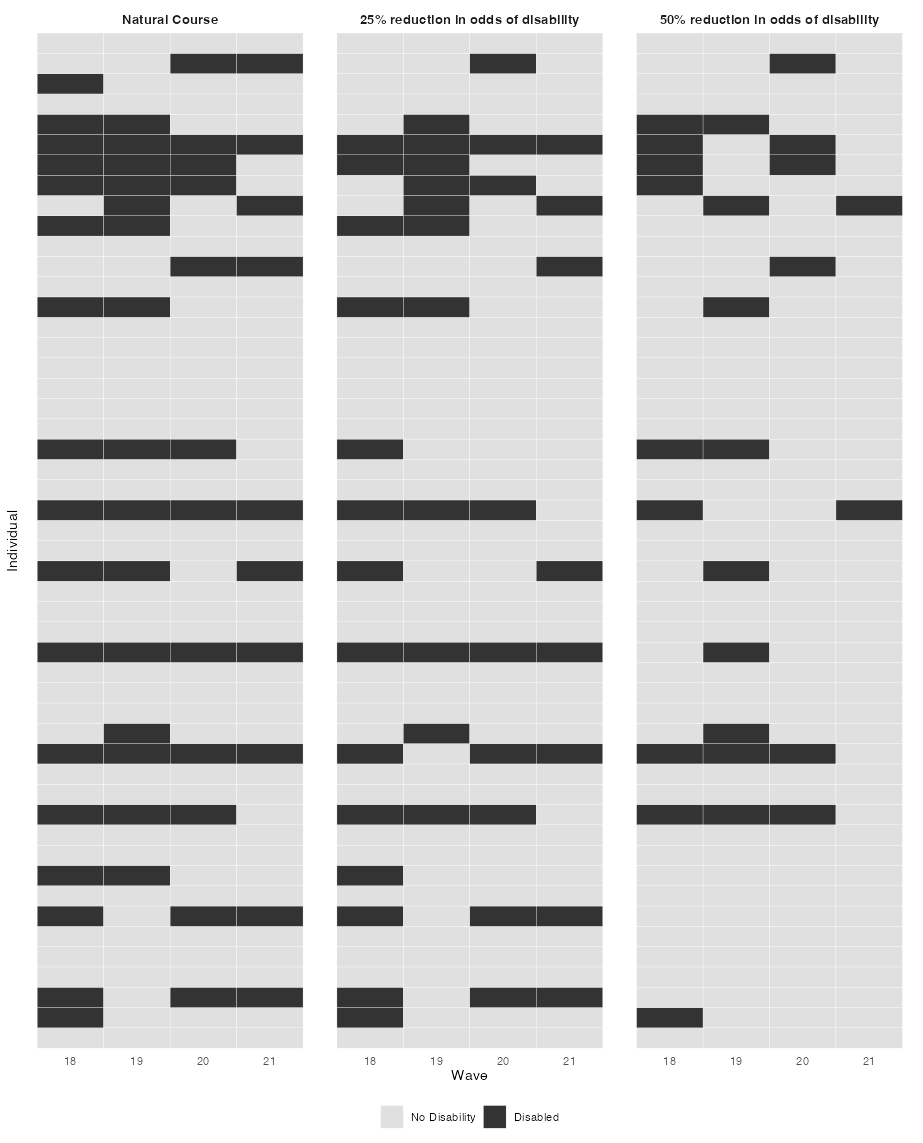

*Note: IPSI indicates incremental propensity score intervention. Dark tiles indicate disability; light tiles indicate no disability.*

**eTable S1. Flow of participant numbers after each follow-up wave in the main wave 18 to 22 analysis.**

| **Wave** | **Retained, n** | **Retained, %** | **Censored since previous wave, n** | **Censored since previous wave, %** |
| --- | --- | --- | --- | --- |
| 18 | 9,635 | 100.0 | - | - |
| 19 | 9,570 | 99.3 | 65 | 0.7 |
| 20 | 9,519 | 98.8 | 51 | 0.5 |
| 21 | 9,479 | 98.4 | 40 | 0.4 |
| 22 | 9,426 | 97.8 | 53 | 0.6 |

*Note. n indicates number of participants. Wave 18 is the baseline wave; not applicable indicates no previous wave for calculating censoring or loss to follow-up.*

**eFigure S2. Distribution of missingness across study variables by covariate and wave before multiple imputation.**

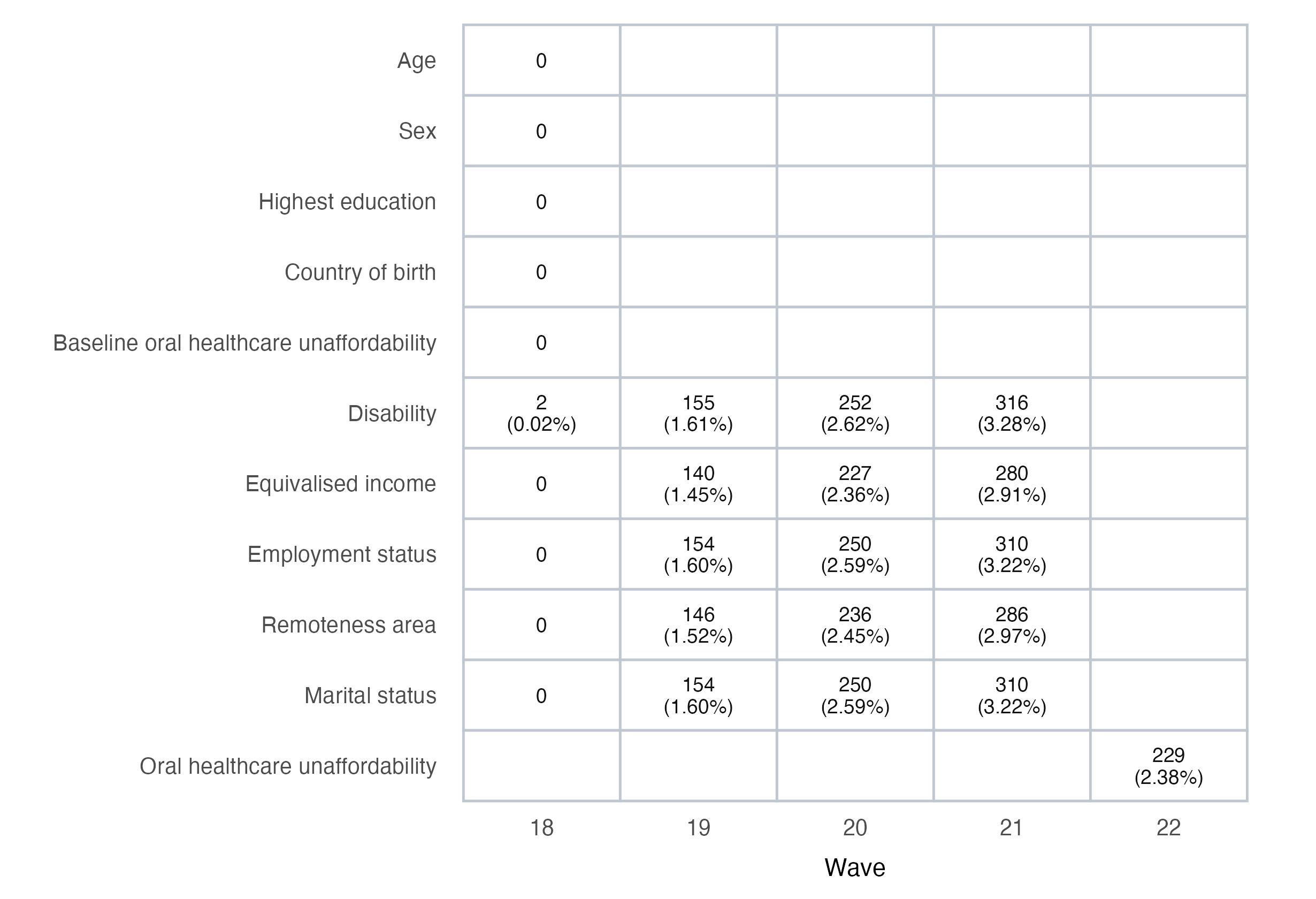

*Note: Values are number missing and percentage missing among main analytic participants before imputation; blank cells indicate variables not included at that wave.*

**eTable S2. Results of the incidence cohort analysis restricted to participants without baseline oral healthcare unaffordability.**

| **Intervention scenario** | **RR (95% CI)** | **RD (95% CI)** |
| --- | --- | --- |
| Sustained disability at every exposure wave | 1.43 (0.89, 2.30) | 0.014 (-0.009, 0.037) |
| No disability at any exposure wave | 0.52 (0.40, 0.67) | -0.016 (-0.021, -0.011) |
| 50% reduction in odds of disability (IPSI) | 0.71 (0.63, 0.81) | -0.009 (-0.013, -0.006) |
| 25% reduction in odds of disability (IPSI) | 0.85 (0.79, 0.91) | -0.005 (-0.007, -0.003) |

*Note: CI indicates confidence interval; IPSI, incremental propensity score intervention; RD, risk difference; RR, risk ratio. Estimates are contrasts with the natural course. The incidence cohort excludes participants who reported oral healthcare unaffordability at baseline.*

**eTable S3. Results of the pre-pandemic waves 14 to 18 analysis.**

| **Intervention scenario** | **RR (95% CI)** | **RD (95% CI)** |
| --- | --- | --- |
| Sustained disability at every exposure wave | 1.98 (1.51, 2.60) | 0.058 (0.025, 0.092) |
| No disability at any exposure wave | 0.70 (0.60, 0.82) | -0.018 (-0.025, -0.011) |
| 50% reduction in odds of disability (IPSI) | 0.78 (0.72, 0.85) | -0.013 (-0.017, -0.009) |
| 25% reduction in odds of disability (IPSI) | 0.86 (0.82, 0.91) | -0.008 (-0.011, -0.005) |

*Note: CI indicates confidence interval; IPSI, incremental propensity score intervention; RD, risk difference; RR, risk ratio. Estimates are contrasts with the natural course. The pre-pandemic analysis uses waves 14 to 18.*
